# The performance of AlphaMissense to identify genes causing disease

**DOI:** 10.1101/2024.03.05.24303647

**Authors:** Yiheng Chen, Guillaume Butler-Laporte, Kevin Y. H. Liang, Yann Ilboudo, Summaira Yasmeen, Takayoshi Sasako, Claudia Langenberg, Celia M.T. Greenwood, J Brent Richards

**Affiliations:** Department of Human Genetics, McGill University, Montréal, QC, Canada; Lady Davis Institute, Jewish General Hospital, McGill University, Montréal, QC, Canada; Department of Epidemiology, Biostatistics and Occupational Health, McGill University, Montréal, QC, Canada; Quantitative Life Sciences Program, McGill University, Montreal, Quebec, Canada; Computational Medicine, Berlin Institute of Health at Charité—Universitätsmedizin Berlin, Berlin, Germany; Tanaka Diabetes Clinic Omiya, Saitama, Japan; Precision Healthcare University Research Institute, Queen Mary University of London, London, UK; MRC Epidemiology Unit, University of Cambridge, Cambridge, UK; Gerald Bronfman Department of Oncology, McGill University, Montreal, Quebec, Canada; 5 Prime Sciences Inc, Montréal, Quebec, Canada; Department of Medicine, McGill University, Montréal, Quebec, Canada; Department of Twin Research, King’s College London, London, UK

## Abstract

A novel algorithm, AlphaMissense, has been shown to have an improved ability to predict the pathogenicity of rare missense genetic variants. However, it is not known whether AlphaMissense improves the ability of gene-based testing to identify disease-causing genes. Using whole-exome sequencing data from the UK Biobank, we compared gene-based association analysis strategies including sets of deleterious variants: predicted loss-of-function (pLoF) variants only, pLoF plus AlphaMissense pathogenic variants, pLoF with missense variants predicted to be deleterious by any of five commonly utilized annotation methods (Missense (1/5)) or only variants predicted to be deleterious by all five methods (Missense (5/5)). We measured performance to identify 519 previously identified positive control genes, which can cause Mendelian diseases, or are the targets of successfully developed medicines. These strategies identified 850k pLoF variants and 5 million deleterious missense variants, including 22k likely pathogenic missense variants identified exclusively by AlphaMissense. The gene-based association tests found 608 significant gene associations (at *P*<1.25×10^−7^) across 24 common traits and diseases. Compared to pLOFs plus Missense (5/5), tests using pLoFs and AlphaMissense variants found slightly more significant gene-disease and gene-trait associations, albeit with a marginally lower proportion of positive control genes. Nevertheless, their overall performance was similar. Merging AlphaMissense with Missense (5/5), whether through their intersection or union, did not yield any further enhancement in performance. In summary, employing AlphaMissense to select deleterious variants for gene-based testing did not improve the ability to identify genes that are known to cause disease.

## Introduction

Rare genetic variants are important contributors to human diseases. They contribute to most Mendelian disorders, and their effect sizes upon common diseases are larger than those attributed to common variants (1–4). Importantly, associated rare genetic variants are often coding and can therefore be directly attributed to a gene. Loss of function rare variants can offer insights into the direction of genetic effect on disease outcome. However, studying rare causal variants is challenging, since most of the genetic variation in the genome is both rare and benign. Thus, gene-based analysis is usually employed to improve statistical power by aggregating multiple rare variants across a gene into one test to improve statistical power to detect disease associations (5).

Previous gene-based multi-variant tests like exome-wide association studies (ExWAS) have successfully identified disease-causal genes, like *WNT1* for osteoporosis (6), and drug-targeting genes, such as *PCSK9* for low-density lipoprotein (LDL)-cholesterol levels (7). Nevertheless, the power of ExWAS relies heavily on the prior identification of variants with a likely functional impact (5) to reduce the number of irrelevant genetic variants included in the tests. While predicted loss-of-function (pLoFs) rare variants are most likely to contribute to gene-based tests, deleterious missense variants can also increase statistical power as they tend to be more common. However, to use deleterious missense variants, one must understand which of the missense variants is most likely to influence protein function—a process referred to as variant annotation. Moreover, all deleterious missense variant annotation strategies must strike a balance between false positive and false negative identification of such variants (8,9).

Recent advances in missense variant effect prediction have made progress towards resolving this problem. AlphaMissense, a recently described method based on an unsupervised language model, combines protein structural context with evolutionary conservation and has claimed to achieve over 90% precision when predicting the known clinical impact of missense variants (9). Additionally, their variant pathogenicity annotations improved the prediction of gene essentiality for cell survival and fitness.

However, it is not known whether the improvements observed in AlphaMissense’s ability to predict the deleteriousness of missense variants results in improved association testing between genes and diseases. If this improvement were striking, it could help to identify new causes of disease and consequently drug targets for needed drug development. Using the UK Biobank whole exome sequencing (WES) data, we tested the ability of AlphaMissense variant annotation to improve the ability to identify positive control genes (known to cause disease) through collapsing gene-based tests on 12 continuous traits and 12 diseases. We compared its performance to other leading algorithms. The results empirically test the ability of AlphaMissense to improve the identification of genes causing disease.

## Methods

### UK Biobank cohort

The UK Biobank is a cohort study that has recruited over 500,000 participants between 40 and 69 years of age at 22 testing centers across the United Kingdom and collected a large set of phenotypes and biological samples. We included in our analyses a total of 444,072 genetically predicted European genetic ancestry individuals with available WES data generated following the OQFE protocol (10) and with measurements of selected phenotypes and diseases. The detailed steps for the sample preparation, sequencing, filtering, and calling of UK Biobank WES data have been previously described (10,11).

### Phenotype definitions

From the UK Biobank, we selected 12 continuous traits and 12 diseases for analysis based on the trait sample sizes and whether there were known disease causal genes or drug target genes for each trait. The continuous traits included estimated bone mineral density, serum triglyceride levels, systolic blood pressure, diastolic blood pressure, standing height, serum low-density lipoproteins, serum bilirubin, serum glucose, red blood cell counts, and serum calcium level, body mass index, and waist-hip circumference ratio and the 12 diseases included hypertension, hypercholesterolemia, diaphragmatic hernia, osteoarthritis (localized), cataract, type 2 diabetes, major depressive disorder, hypothyroidism, acute renal failure, atrial fibrillation, cancer of prostate (males only) and breast cancer (females only). The sample sizes for the analysis of each trait and disease can be found in Supplementary table 1. ICD-10 codes were grouped to construct diseases following the phecodes system (12). The list of used ICD-10 codes for each phecode can be found in Supplementary table 2.

### Variant annotation

We annotated the variants from exome sequencing after alignment using the Ensembl Variant Effect Predictor (VEP) (v.110). Variant annotations among transcript ablation, splice acceptor, splice donor, stop gained, frameshift, stop lost, start lost, transcript amplification, feature elongation, and feature truncation, were considered as predicted loss-of-function (pLoF) variants (10). Missense variants were classified with two strategies. The first strategy used AlphaMissense (9), and missense variants were included in our analyses if AlphaMissense predicted them to be “likely pathogenic”. We built a second strategy by combining results from five commonly used annotation methods (i.e., SIFT (13), PolyPhen2 (HDIV) (14), PolyPhen2 (HVAR) (15), MutationTaster (16), and LRT (17)). We classified a missense variant as “likely deleterious” if all five algorithms predicted it to be deleterious (i.e., Missense (5/5)), and “possibly deleterious” if at least one of the five algorithms predicted it to be deleterious (i.e., Missense (1/5)), similar to methods used before (6,10).

### Gene-based disease and trait association test

For each gene, variant annotations and alternative allele frequency (AAF) categorized the inclusion of variants into 20 gene burden exposures, created by the combination of four annotation mask definitions and five AAF thresholds and statistical testing method combinations. The four masks categories included: (1) pLoF variants; (2) pLoF or “likely pathogenic” variants by AlphaMissense (pLoF with AlphaMissense); (3) pLoF or “likely deleterious” missense variants by the five commonly used methods (pLoF with Missense (5/5)); (4) pLoF or “possibly deleterious” missense variants by the five commonly used methods (pLoF with Missense (1/5)). The five AAF and statistical test method combinations included (1) standard burden test with AAF < 1%; (2) standard burden test with AAF < 0.1%; (3) standard burden test with singletons; (4) SKAT variance-component test with AAF <1%; (5) SKAT-O combined test with AAF <1%). The smallest p-value of the five AAF and test combinations for each gene under different masks were retained for subsequent significance and classification testing. For our primary method, we built masks for burden tests using the maximum number of alternative alleles found across all selected variant sites of a gene. As a sensitivity analysis, we also tested whether building masks by total number of alternative alleles across these sites, a approach assuming these sites have cumulative effect, would impact the results of association analyses.

All analyses were performed using Regenie software (18). The regression analyses included age, age^2^, sex, sex*age, sex*age^2^, 10 genetic principal components (PC) obtained from common genetic variants (MAF>1%), and 20 genetic PCs obtained from rare genetic variants (MAF<1%) as covariates. The statistical significance threshold was *P* < 1.25×10^−7^ (0.05 / (approximately 20,000 genes * 20 gene-burden exposures)).

### Selection of positive control genes

To evaluate whether different masks have different abilities to identify genes that were known to cause Mendelian forms of disease, or the targets of successfully developed medicines, we compiled a list of positive control genes from two sources. We first included positive control genes from two previous studies where these genes were used to train their algorithms to prioritize disease-causal or drug-targeting genes from genome-wide association study (GWAS) signals (19,20). Their positive control gene lists were generated by combining genetic evidence, drug–target–indication associations, and manual curation from board certified physicians and domain experts. Additionally, we included Mendelian diseases genes from the MendelVar database which was created by integrating functional annotations from the Online Mendelian Inheritance in Man (OMIM), Deciphering Developmental Disorders Study (DECIPHER), Orphanet and Genomics England databases (21). The full list of 509 positive control genes for the selected traits and diseases can be found in Supplementary Table 3.

### Evaluation of classification accuracy

The ability to accurately identify positive control genes using gene burden tests with different variant sets and mask settings was measured by the area under the receiver-operator curves (AUROC) and precision-recall curves (AUPRC). Specifically, PRC and ROC were generated using results from 21 traits and diseases where we could confirm at least one positive control gene. The 95% confidence intervals (CI) for AUROC and AUPRC were determined using 1000 bootstrap replicates. The baseline for AUROC is 0.5, an uninformative classifier. The baseline level for AUPRC is 0.0018 which equals the proportion of positive control genes among tested genes.

## Results

Starting with 19,606 genes, for every exon, we annotated deleterious variants into four categories: pLoF, AlphaMissense, Missense (5/5), and Missense (1/5). Then we assembled four sets of predicted deleterious variants (i.e., masks): (1) pLoF, (2) pLoF with AlphaMissense, (3) pLoF with Missense (5/5), and (4) pLoF with Missense (1/5). Each mask provided a list of variants for genes in gene-based association analysis. Lastly, we retained the smallest p-values from the five different combinations of alternative allele frequency and statistical test method for the association between each gene and each tested trait or disease under different masks (Figure 1a).

**Figure 1.**
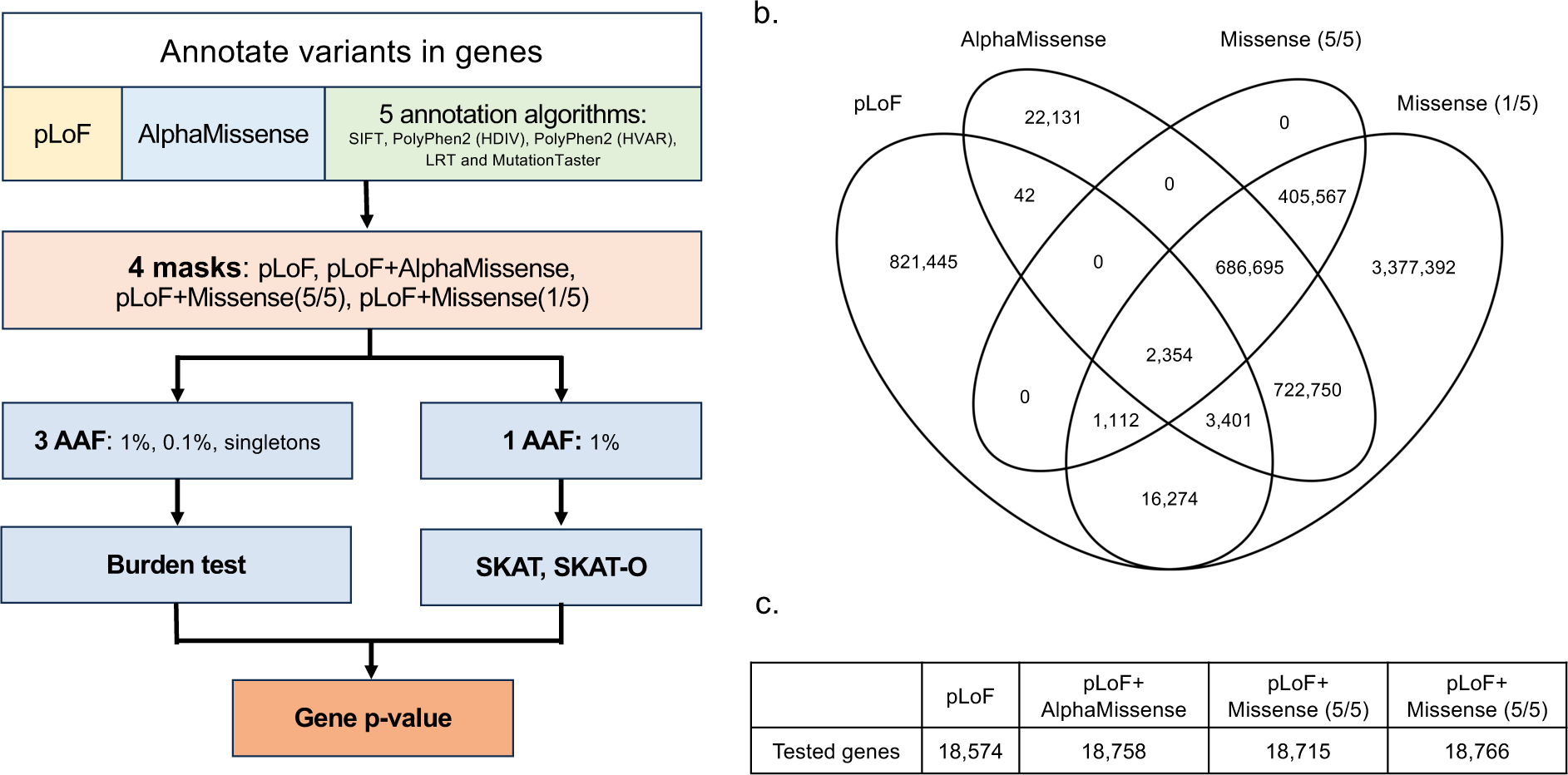
Overview of ExWAS analysis and comparison of variants annotations by different methods. (a) strategy to obtain single *P* value for each gene (b) overlap of annotated likely deleterious variants by different ExWAS mask settings (c) Number of tested genes with different mask settings.

Of 26 million variants from UK Biobank WES data, we identified 850k pLoF variants and 5 million predicted deleterious missense variants by AlphaMissense or any of the five commonly used annotation methods (i.e., SIFT, PolyPhen2 (HDIV), PolyPhen2 (HVAR), MutationTaster, and LRT). Specifically, AlphaMissense classified 1.4 million variants as “likely pathogenic”, including 22k identified exclusively by AlphaMissense. Missense (1/5) captured over 98% of AlphaMissense predicted “likely pathogenic” variants while Missense (5/5) covered 48% of AlphaMissense predicted “likely pathogenic” variants (Figure 1b). Moreover, our results showed that among the masks evaluated, Missense (1/5) labeled the highest number of deleterious variants per gene on average (267 variants per gene), followed by AlphaMissense (74 variants per gene), Missense (5/5) (56 variants per gene), and pLoF (43 variants per gene) (Supplementary Table 4). Despite the considerable variance in the number of annotated variants across different annotation categories, 99% of genes were tested in all masks (Figure 1c).

In the exome-wide gene-based analysis, we first checked the genomic inflation factors of the p-values for each mask and test method combination. In general, no strong genomic inflation was observed (value range: 0.96-1.37) except for standing height (value range: 1.11-1.94) (Supplementary Table 5). This is not surprising as height is a well-known highly polygenic trait (22).

In total, our gene-based association tests found 608 significant gene associations (*P* <1.25×10^−7^) across 24 common traits and diseases. We found that adding predicted deleterious missense variants to masks led to the identification of at least 60% more significant gene-trait associations and about 30% more positive control genes as compared to pLoF-only mask (Figure 2, Supplementary Figure 1a, and Supplementary Table 6). Despite different numbers of associations identified, 114 significant associations and 30 positive control genes were captured using any of the masks, which accounts for between 27-57% and 50-71% of the findings, respectively, of each mask (Supplementary Figure 1b and Supplementary Figure 1c). Comparing across four masks, pLoF with AlphaMissense and pLoF with Missense (5/5) resulted in more significant associations and positive control genes than the pLoF-only mask while keeping a lower false positive rate than pLoF with Missense (1/5) mask, indicating their superiority over the other masks. Between these two preferred masks, pLoF with AlphaMissense identified largely similar or slightly higher numbers of significant gene-trait and gene-disease associations compared to pLoF with Missense (5/5). Meanwhile, these two methods demonstrated a similar sensitivity in capturing positive control genes, as indicated by the proportions of positive control genes among significant associations (17.9% for pLoF with Missense (5/5), and 17.5% for pLoF with AlphaMissense) (Figure 2). Furthermore, the pLoF with AlphaMissense and pLoF with Missense (5/5) masks shared 245 (71 and 75%) significant association findings and 46 (77 and 80%) of identified positive control genes (Supplementary Figure 1b and 1c).

**Figure 2.**
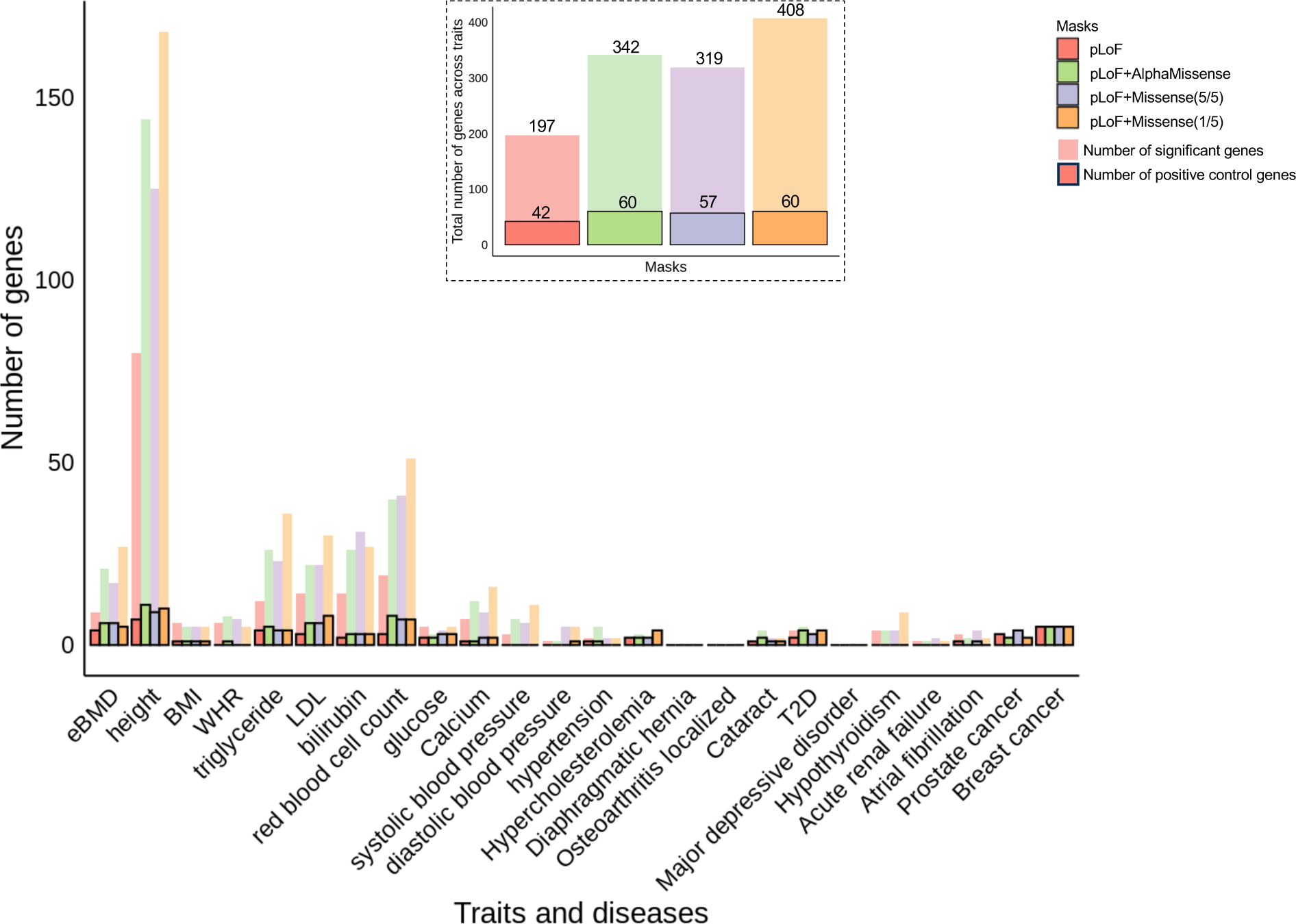
Significant gene associations identified in exome-wide gene burden analysis across 12 traits and 12 diseases. The bars without outlines indicate the numbers of significant genes (*P* < 1.25×10^−7^) identified in each trait and disease by different masks. The bars with outlines indicate the number of significant genes that are also positive control genes for each trait and diseases identified by different masks. The inset figure shows the total number of significant genes and positive controls identified by each mask across all the tested traits and diseases. Abbreviations: estimated bone mineral density (eBMD), body mass index (BMI), waist-hip circumference ratio (WHR), serum low-density lipoproteins (LDL), type 2 diabetes (T2D).

Next, to evaluate whether different masks enhanced the distinction between positive control genes and non-positive control genes by offering more divergent *P* values, we evaluated the performance of using different masks in classifying these genes by calculating the operating characteristic curve (ROC) and precision-recall curve (PRC). Upon comparison, we observed that all four masks have statistically indistinguishable area under the receiver-operator curves (AUROC) (Figure 3, left panel). However, pLoF with Missense (5/5) and pLoF with AlphaMissense have a higher estimated area under the precision-recall curves (AUPRC) than the other two masks despite the fact that all the 95% confidence intervals of AUPRCs overlapped (Figure 3, right panel). Similar AUROC and AUPRC patterns can be observed across tested traits, but we did observe that specific masks could perform better for certain traits and diseases (Supplementary Figure 2). Additionally, we tested whether using different aggregating methods for counting alleles, in burden tests, across genetic sites within genes changed the mask performance. Using the maximum number of alternative alleles across sites (the default approach) and using the sum of the number of alternative alleles in gene-based association analyses performed similarly (Supplementary Figure 3).

**Figure 3.**
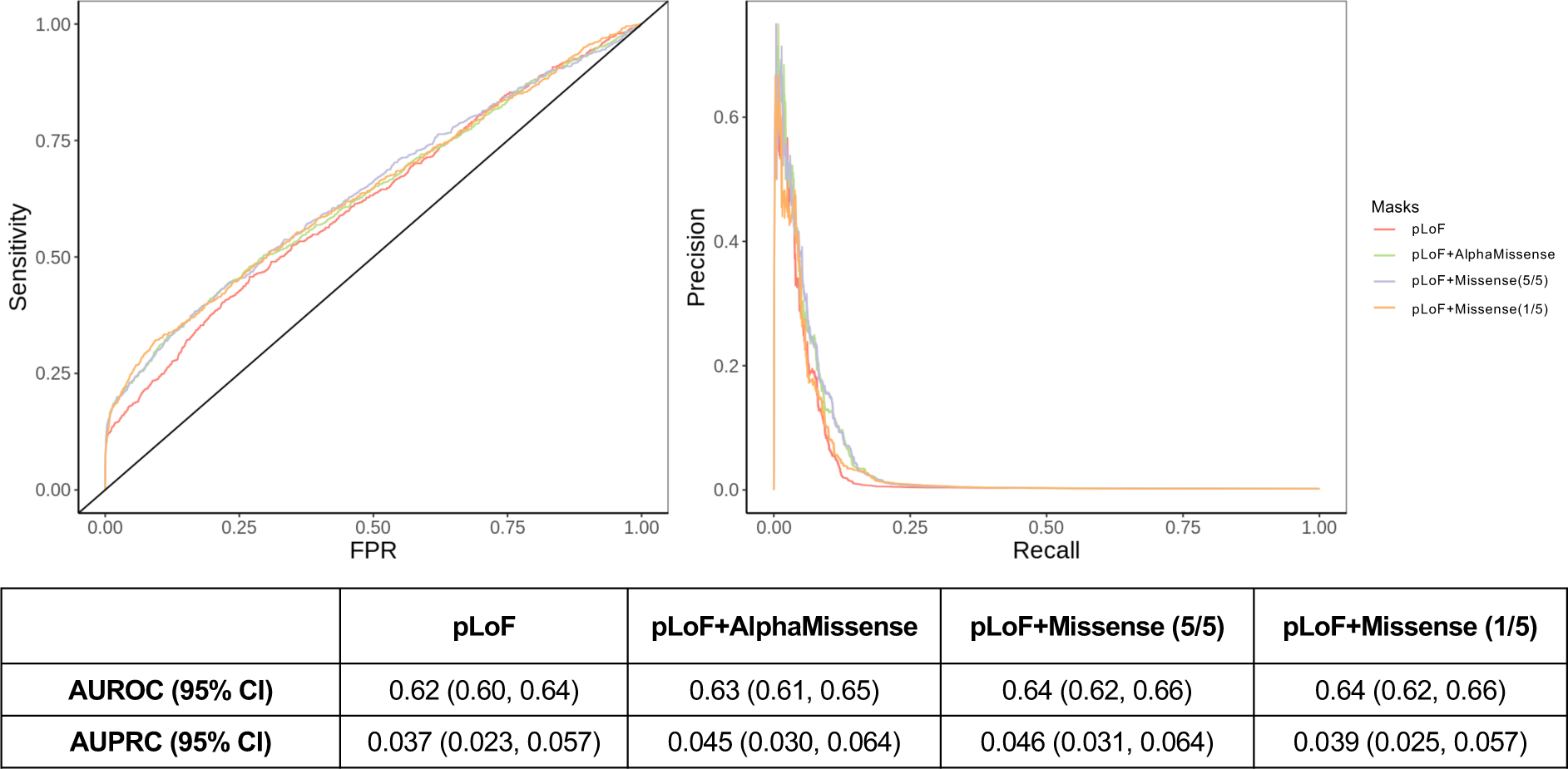
Performance curves (ROC and PRC) for all four masks to identify positive controls genes across all 21 tested traits and diseases with positive control genes.

Considering that performance was better when pLoF variants were combined with either Missense (5/5) or AlphaMissense annotated deleterious variants, we further investigated whether merging AlphaMissense and Missense (5/5) annotations before combining with the pLoF variants could improve their ability to classify positive control genes. We tested two designs: using pLoF variants and variants predicted to be deleterious by (1) both AlphaMissense and Missense (5/5) or by (2) either AlphaMissense and Missense (5/5).

As shown in Figure 4, utilizing deleterious variants predicted by either method identified slightly more significant associations (372 pairs), although the precision remained similar (17.7%) (Supplementary Table 7). In contrast, using the overlapping predictions led to fewer significant associations (287 pairs) but marginally higher precision (18.5%). The AUROC and AUPRC of these two new mask definitions are similar to other masks (Supplementary Figure 4). Overall, little improvement was observed by merging Missense (5/5) with AlphaMissense.

**Figure 4.**
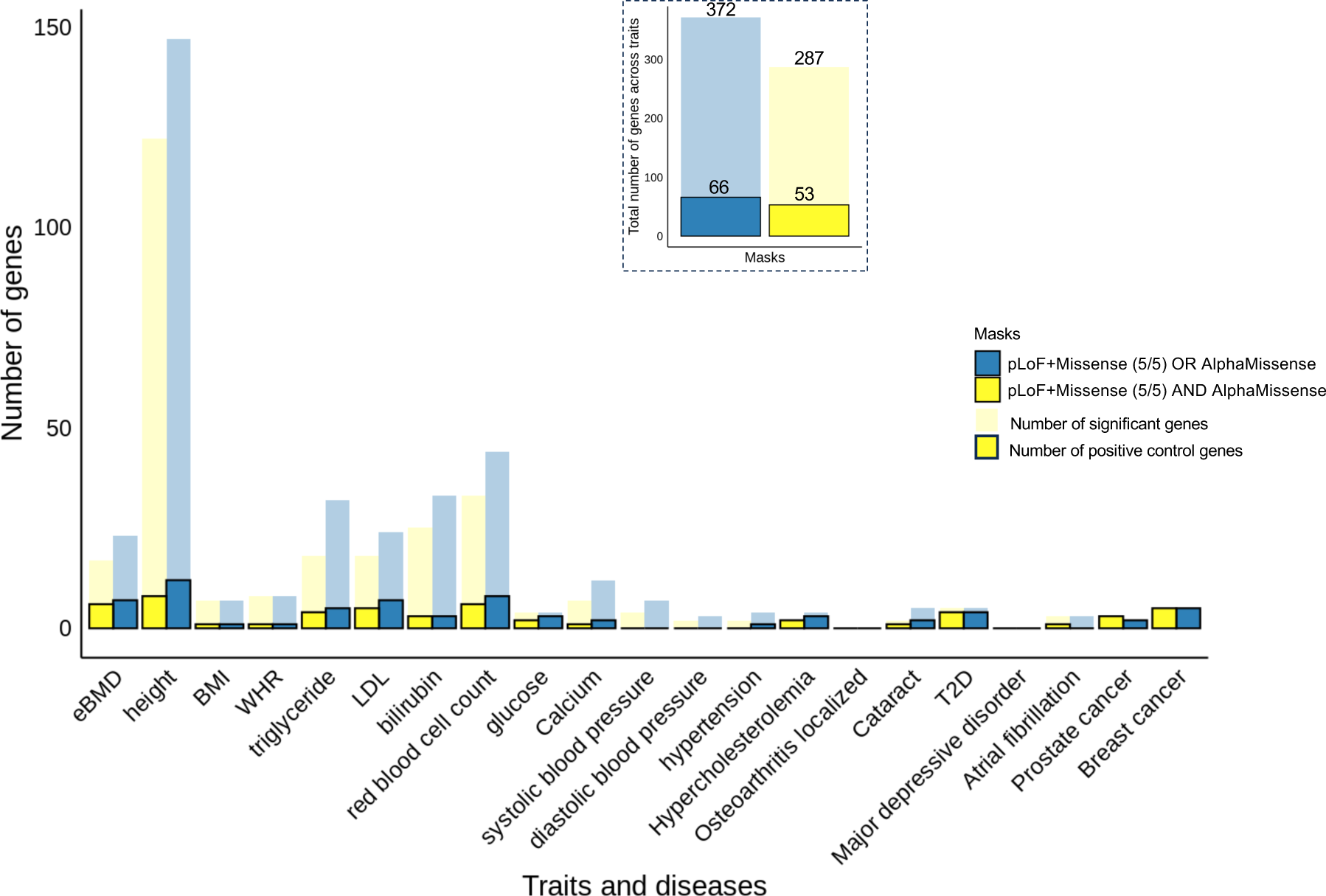
Significant gene-trait and gene-disease associations identified in exome-wide gene burden analysis across 24 traits using pLoF with the intersection or union of predicted deleterious variants by AlphaMissense and Missense (5/5). The inset figure shows the total number of significant genes and positive controls identified by each mask across all the tested traits and diseases. Abbreviations: estimated bone mineral density (eBMD), body mass index (BMI), waist-hip circumference ratio (WHR), serum low-density lipoproteins (LDL), type 2 diabetes (T2D).

## Discussion

Gene-based tests offer an elegant way to study the effect of rare coding variants on human traits by improving statistical power. However, the best way to combine genetic variants into gene sets is still not fully determined, simply because there are usually many irrelevant genetic variants in each gene set which may dilute any signal from the set of causal variants. Hence, such analyses usually rely on algorithms to predict which variants are likely to be loss of function or missense variants with deleterious effects. As gene-based analyses are restricted to a likely deleterious subset of variants to increase this signal to noise ratio, the success of these analyses rest partially on the performance of the predictions. The emergence of a language model-based variant effect prediction methods, AlphaMissense, has been suggested to be able to improve gene-based association. However, our results showed that AlphaMissense did not importantly outperform the current state-of-the-art masks in gene-based association analyses using whole-exome data.

There are multiple reasons why the inclusion of ‘likely pathogenic’ missense variants, as annotated by AlphaMissense, does not lead to significant improvements. First, the masks used in our analysis always included pLoF variants, which already contribute significantly to the associations observed between genes and traits. Furthermore, the addition of AlphaMissense’s predicted pathogenic missense variants expands the analyzed gene pool by only 184 genes (when added to pLOFs) or 33 genes (when added to pLOF and Missense (5/5)) beyond those tested using pLoF-only masks. This modest increase in the number of genes tested offers limited scope for enhancing the performance of gene-based association tests. Lastly, as noted earlier in this report, other missense annotation methods largely capture the same ‘likely pathogenic’ variants identified by AlphaMissense. Given that all gene-based tests then summarize information across all analyzed variants in a gene (in various ways), the small number of differently-prediction variants may not render a large difference in the associated genes.

AlphaMissense may provide useful and clarifying information in scenarios where understanding single variant effects is crucial. For example, AlphaMissense could be particularly helpful in pinpointing actionable genetic sites within known disease-causing genes. This may be particularly useful for patients with Mendelian diseases without major structural disruptions in the genetic region (23,24). Additionally, since AlphaMissense integrates protein structure context into its predictions of variant effects, it should be more effective when identifying deleterious variants for diseases where protein malfunction arises from changes in protein conformation. AlphaMissense could also be advantageous in predicting pharmacogenetic effects that involve protein-drug interactions (25).

We recognize that while pLoF and missense variant annotations should not be affected by genetic ancestry, we only performed our analyses in European genetic ancestry individuals from the UK Biobank, and we only examined 24 traits. Hence, these results will need replication in other populations once sample sizes allow this. Second, the UK Biobank cohort is a relatively healthy cohort. The number of disease cases is low, which can limit the statistical power to identify disease-related genes, which may make it more difficult to compare the performance of different masks in ExWAS. Lastly, there are other annotation masks that we have not tested, and which may perform differently. Nevertheless, we compared our results to the best currently available annotations (10), and we have established that any future work should make comparisons to AlphaMissense.

In summary, we found that most of the “likely pathogenic” missense variants identified by AlphaMissense were also generally predicted to be deleterious by at least one of five commonly used variant annotation methods. Using masks combining AlphaMissense with pLoF did not outperform the state-of-the-art missense annotation tools for gene-based studies.

## Data availability

Individual-level genotype, exome sequencing, and phenotype data is available to approved researchers via UK Biobank at: https://www.ukbiobank.ac.uk. ExWAS summary statistics will be made available at GWAS Catalog (https://www.ebi.ac.uk/gwas/).

## Code availability

VEP software can be downloaded at https://github.com/Ensembl/ensembl-vep. Regenie software can be found at https://github.com/rgcgithub/regenie. UK Biobank exome data was analyzed using Regenie 3.2.1. All other data analysis was performed using R (v.4.1.2). Additional codes can be accessed through Github upon publication.

## Acknowledgments

We appreciate the individuals who participated in UK Biobank. This research has been conducted using UK Biobank data under application ID 27449.

The Richards research group is supported by the Canadian Institutes of Health Research (CIHR: 365825, 409511, 100558, 169303), the McGill Interdisciplinary Initiative in Infection and Immunity (MI4), the Lady Davis Institute of the Jewish General Hospital, the Jewish General Hospital Foundation, the Canadian Foundation for Innovation, the NIH Foundation, Cancer Research UK, Genome Québec, the Public Health Agency of Canada, McGill University, Cancer Research UK, and the Fonds de Recherche Québec Santé (FRQS). J.B.R. is supported by an FRQS Mérite Clinical Research Scholarship. Support from Calcul Québec and Compute Canada is acknowledged. TwinsUK is funded by the Welcome Trust, Medical Research Council, European Union, the National Institute for Health Research (NIHR)-funded BioResource, Clinical Research Facility and Biomedical Research Centre based at Guy’s and St Thomas’ NHS Foundation Trust in partnership with King’s College London. Y.C. is supported by an FRQS doctoral training fellowship and the Lady Davis Institute/TD Bank Studentship Award. G.B.L. is supported by scholarships from the FRQS, the CIHR, and Québec’s ministry of health and social services.

## Author Contribution Statement

YC - Writing initial draft. YC, GBL, KYHL, CMTG, JBR – Methodology. YC, GBL, KYHL - Data Analysis. YC, GBL, KYHL, YI, SY, TS, CL, CMTG, JBR - Writing review and editing draft. JBR – Supervision. All authors commented/revised the manuscript and agreed to its final submitted version.

## Ethical approval

The UK Biobank was approved by the North West Multi-centre Research Ethics Committee and informed consent was obtained from all participants prior to participation.

