## Supplementary Figures for "The performance of AlphaMissense to identify genes causing disease"

**Supplementary Figure 1.** Comparison of the significant and positive control gene-trait and gene-disease pairs identified by ExWAS using different masks. (a) Upset plot shows the degree of overlap between identified significant genes and those designated as positive control genes (b) Venn diagram for significant gene-trait or gene-disease associations identified by different masks (c) Venn diagram for positive control genes identified by different masks.

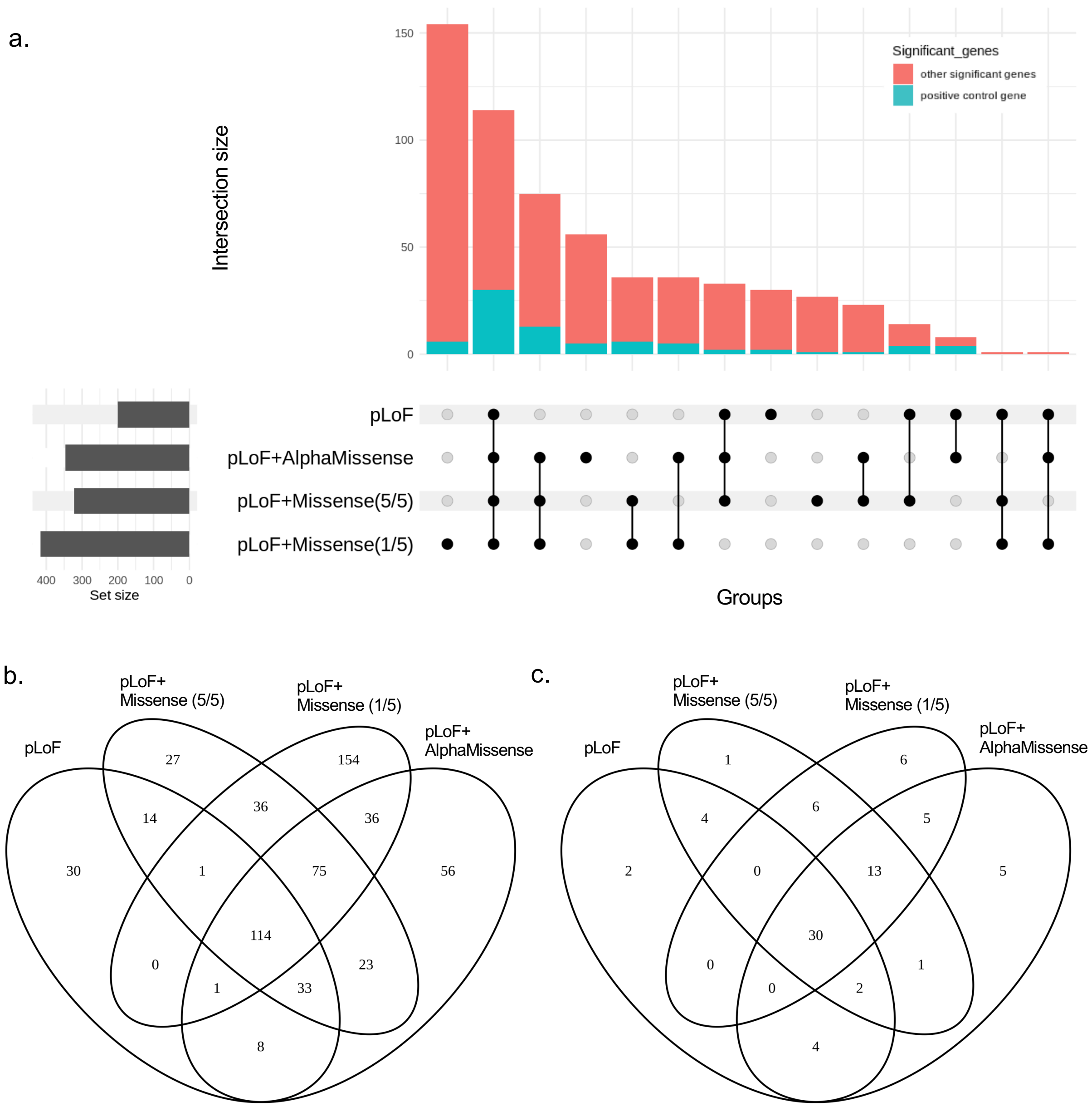

**Supplementary Figure 2.** Performance by AUROC and AUPRC for all four masks on identifying positive controls genes in each of the tested traits and diseases. Abbreviations: estimated bone mineral density (eBMD), body mass index (BMI), waist-hip circumference ratio (WHR), serum low-density lipoproteins (LDL), type 2 diabetes (T2D).

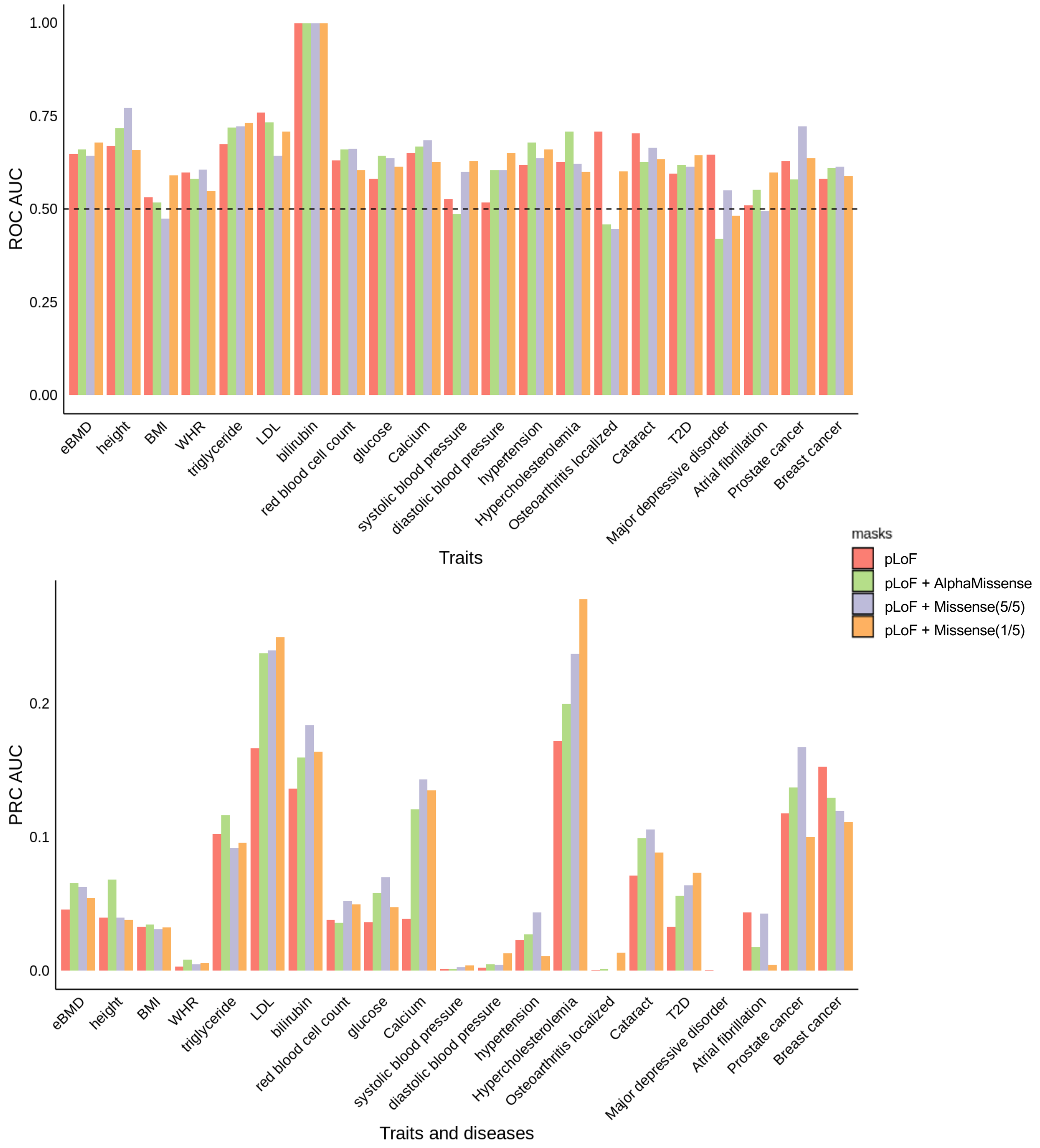

**Supplementary Figure 3.** Performance curves (ROC and PRC) for all four masks on identifying positive controls genes across tested traits and diseases using the sum of alternative alleles to pool deleterious variants.

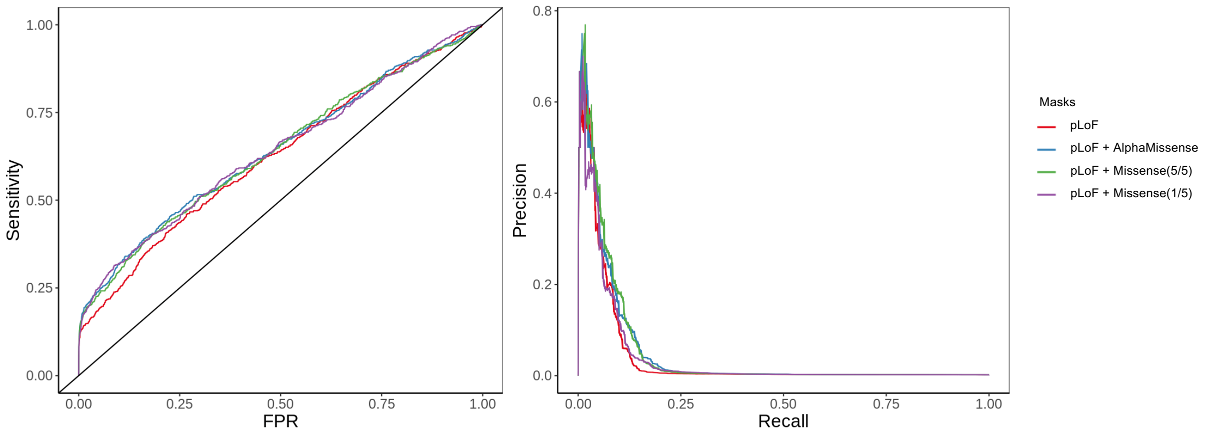


| | pLoF | pLoF+AlphaMissense | pLoF+Missense (5/5) | pLoF+Missense (1/5) |
|---|---|---|---|---|
| **AUROC (95% CI)** | 0.62 (0.60, 0.65) | 0.64 (0.62, 0.67) | 0.64 (0.62, 0.67) | 0.64 (0.62, 0.66) |
| **AUPRC (95% CI)** | 0.040 (0.026, 0.059) | 0.048 (0.030, 0.064) | 0.050 (0.031, 0.070) | 0.041 (0.026, 0.063) |

**Supplementary Figure 4.** Performance curves (ROC and PRC) for identifying positive control genes across tested traits and diseases by six mask settings.

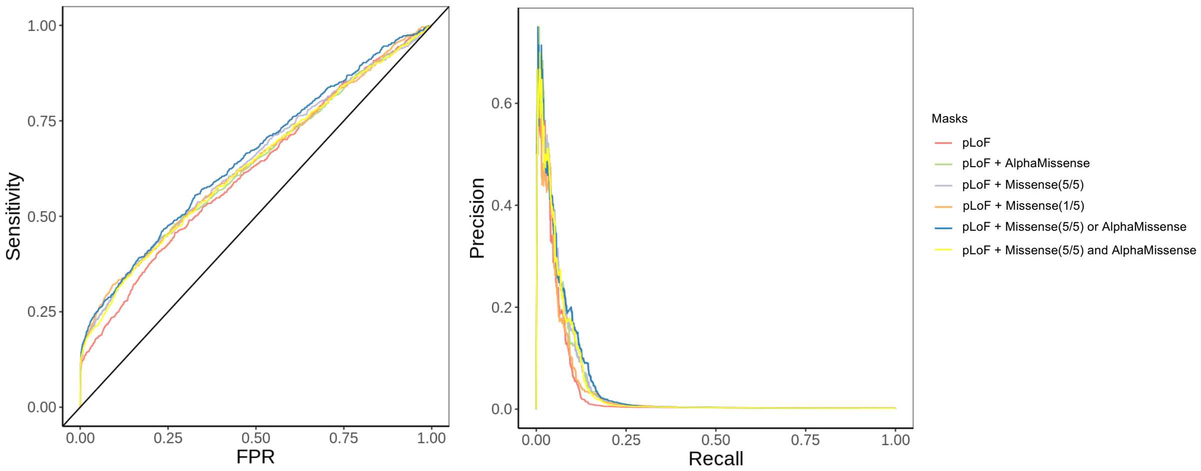


| | **pLoF** | **pLoF+ AlphaMissense** | **pLoF+ Missense (5/5)** | **pLoF+ Missense (1/5)** | **pLoF+ Missense (5/5) OR AlphaMissense** | **pLoF+ Missense (5/5) AND AlphaMissense** |
|---|---|---|---|---|---|---|
| **AUROC (95% CI)** | 0.62 (0.60,0.64) | 0.63 (0.61, 0.65) | 0.64 (0.62,0.66) | 0.64 (0.62,0.66) | 0.66 (0.63, 0.68) | 0.63 (0.61, 0.66) |
| **AUPRC (95% CI)** | 0.037 (0.023, 0.057) | 0.045 (0.030, 0.064) | 0.046 (0.031, 0.064) | 0.039 (0.025, 0.057) | 0.048 (0.033, 0.067) | 0.044 (0.030, 0.063) |
